# Seasonal influenza vaccine hesitancy profiles and determinants among Chinese children’s guardians and the elderly

**DOI:** 10.1101/2021.02.18.21251972

**Authors:** Zheng Wei, Xiu Sun, Yingying Yang, Siyi Zhan, Chuanxi Fu

## Abstract

**Background:** Seasonal influenza imposes a significant clinical and economic burden, yet vaccine coverage remains low in China due to possible influenza vaccine hesitancy (IVH) and practical issues. We sought to investigate IVH and its determinants among children’s guardians and the elderly for better understanding of the situation and for future intervention.

**Methods:** IVH scales for the guardians and elderly were designed. We then performed two cross-sectional studies to identify the profiles and determinants of IVH using stratified cluster random sampling in an eastern China province in 2019.

**Results:** Of the 1,564 guardians and 522 elders, 43.2% (95% confidence interval: 40.4–46.0%) of guardians and 33.5% of elders (95% confidence interval: 29.5–37.6%) had IVH, whereas 31.3% of the guardians and 5.5% of the elderly had Influenza vaccine demand. The scales were well structured (KMO: 0.736 and 0.682; Cronbach’s α: 0.638 and 0.611). ‘From rural area’ (Odds ratio: 1.36), ‘don’t know government recommendation for flu vaccination’ (1.39), ‘don’t know flu vaccine is vaccinated annually’ (1.93), ‘family members (0.22), friends and neighbors had positive attitude toward flu vaccine’(0.58) were related factors of the guardians’ IVH. ‘Aged 70–79 years’ (0.46), ‘had flu before’ (0.35) and ‘once had been vaccinated’ (0.42) were related to the elderly’s IVH.

**Conclusion:** Poor awareness of influenza and vaccination, relatives’ negative/positive attitude, lack of government recommendations, anxiety about vaccine quality, and practical issues such as short supply are related to IVH in China. Precision education aiming hesitancy in wider groups are anticipated to increase vaccine confidence and coverage in influenza-vulnerable groups.

## Introduction

Seasonal influenza, causing 3 to 5 million cases of severe illness and 290 to 650 thousand respiratory deaths annually, with the highest burden in young children and older adults, represents a public health threat with significant socioeconomic implications^1-2^. Seasonal influenza vaccine is moderately effective against the influenza virus infection and more importantly, can reduce intensive care admissions and duration of hospitalizations^3-4^. The World Health Organization (WHO) recommends annual seasonal influenza vaccination for children, the elderly, individuals with underlying health conditions, health-care workers, and pregnant women^5^. Despite disease severity and availability of safe vaccines, low influenza vaccine uptake in specific risk groups remains a challenge to public health wideworld^1,6,7,8^. In China mainland, it is estimated that over 88,000 influenza-associated exorbitant deaths occur annually^1,9^. Although children and the elderly are recommended as prioritized groups for influenza vaccination in China, the coverage is usually low, i.e., 28.4% (95% confidence interval [CI]: 23.6–33.2%) in children aged 6–59 months, and 26.7% (95% CI: 23.8–29.7%) in the elderly^10^.

Vaccine hesitancy (VH) is described as one of the ten threats to global health by WHO in 2019. VH threatens the historical achievements made in reducing the burden of infectious diseases^11^ and fueling the resurgence of vaccine-preventable diseases and plummeting the vaccination rates worldwide^12^. VH has been an important issue of scientific inquiry within high-income countries; vaccine-preventable diseases such as measles have re-emerged in these areas and resulted in substantial consequences for public health and economy^13-14^. In 2014, WHO Strategic Advisory Group of Experts (SAGE) developed the definition and determinants of the Matrix of VH ^15,16^. Data from the WHO/UNICEF Joint Report Form showed that VH had been reported in > 90% of 194 WHO’s member states; meanwhile, the abundant factors covered 22 of the 23 determinants in WHO’s matrix categories, yielding to the complexity of global VH^17^.

In this context, influenza vaccine hesitancy (IVH) can reduce the vaccination coverage against seasonal influenza ^18,19^. With regard to the influenza vaccine, there are traits that are similar to those of other vaccines, which should also be addressed when looking at IVH^18,20^. First, in China, the influenza vaccine is categorized as a non-Expanded Programmed Immunization vaccine, and Chinese residents pay for the vaccination; Second, the knowledge about influenza and vaccination is poor in China, i.e., some people have never even heard of an influenza vaccine ^21,22^; third, insufficient supply of the influenza vaccine from either domestic or international manufacturers cannot meet the increasing demand over the recent years, and thus, it’s common to not seek the influenza vaccination service in Chinese immunization clinics^15^.

Children and elders are both influenza-vulnerable groups, and vaccination is recommended by the Chinese Center for Disease Control and Prevention (CCDC)^3^. However, most IVH studies have been conducted in western and developed countries^18^, and the effect of IVH on vaccination coverage in China still remains unknown. In this study, we investigated the profiles and related determinants of IVH among Chinese children’s guardians and the elders for better understanding of IVH and for future intervention.

## Methods

### VH and vaccine demand

With regard to the following assumptions, VH and vaccine demand were defined in the study: (i) The definitions of VH and vaccine demand are often confusing and need to be clarified; existing term cause problems because of translation discrepancies between different languages^23^; (ii) The impact of VH is continuous, considering that people who received the vaccination in the previous season may refuse it in the current season^18^; (iii) VH has not been defined in China before.

#### IVH

Knowing that the influenza vaccine and vaccination service is available, but one is not completely confident whether to vaccinate, or is still worried after vaccination. It is a state between complete acceptance and rejection. IVH is determined by various influencing factors, and the determinants of IVH for particular populations are different. In the study, we used a five-point Likert question “your future influenza intention to vaccination” to distinguish IVH from the complete acceptance/rejection of the influenza vaccination. Options included: (i) completely reject, (ii) reject but still considering, (iii) have not decided yet or never thought about it, (iv) accept but still considering, (v) completely accept. Respondents who chose option 2, 3, or 4 were considered to have IVH.

#### Influenza vaccine demand (IVD)

Knowing influenza vaccine and vaccination service is available, fully accepting influenza vaccine and actively seeking influenza vaccination service. IVD is included in IVH, that is, people with vaccine demand do not have VH at all, but people without VH may not have the demand. If IVD exceeds vaccine supply, short supply will emerge. In the study, as the future vaccination plan was unknown, we used an item of “influenza vaccination experience” to predict IVD. Those who completely accepted it and had been vaccinated in the previous year (2018/2019 season) were considered to have IVD.

## Participants and design

From July to October 2019, we conducted the cross-sectional design study to collect self-reported data through field survey in eight cities in Zhejiang province. Zhejiang is a mid-developed, east-coastal province of China, with an area of 1,105.5 km^2^ and a population of approximately 70 million peolpe^24^, and there are two influenza epidemics peaks annually, one from January to February and the other from June to August^3^. The stratified cluster random sampling method was used to select eight prefecture-level cities based on geographic location and population composition (Fig 1a). We then randomly selected 2–4 street (township) community health service centers (CHSCs) in each city (Fig 1b).

**Figure 1.**
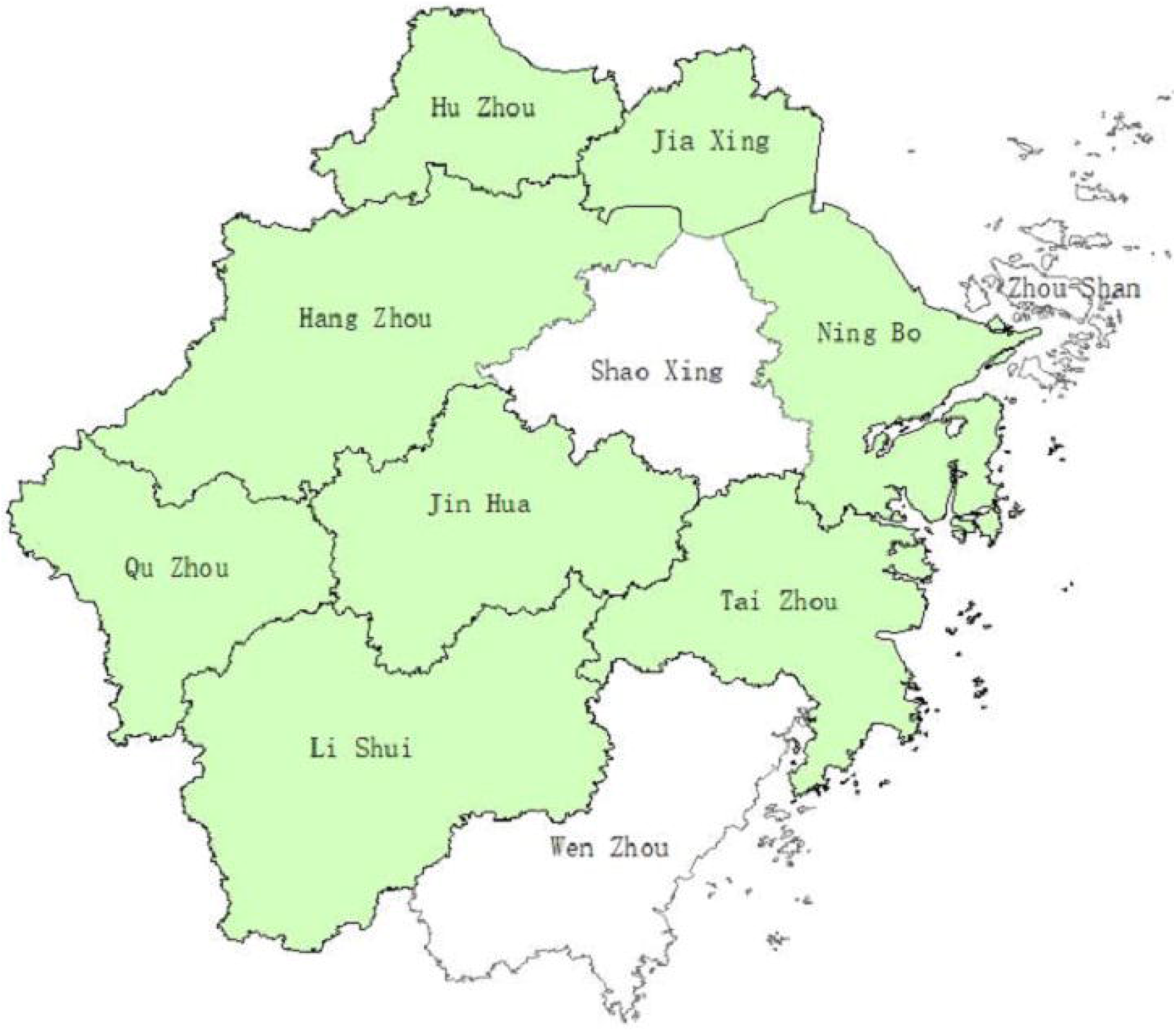
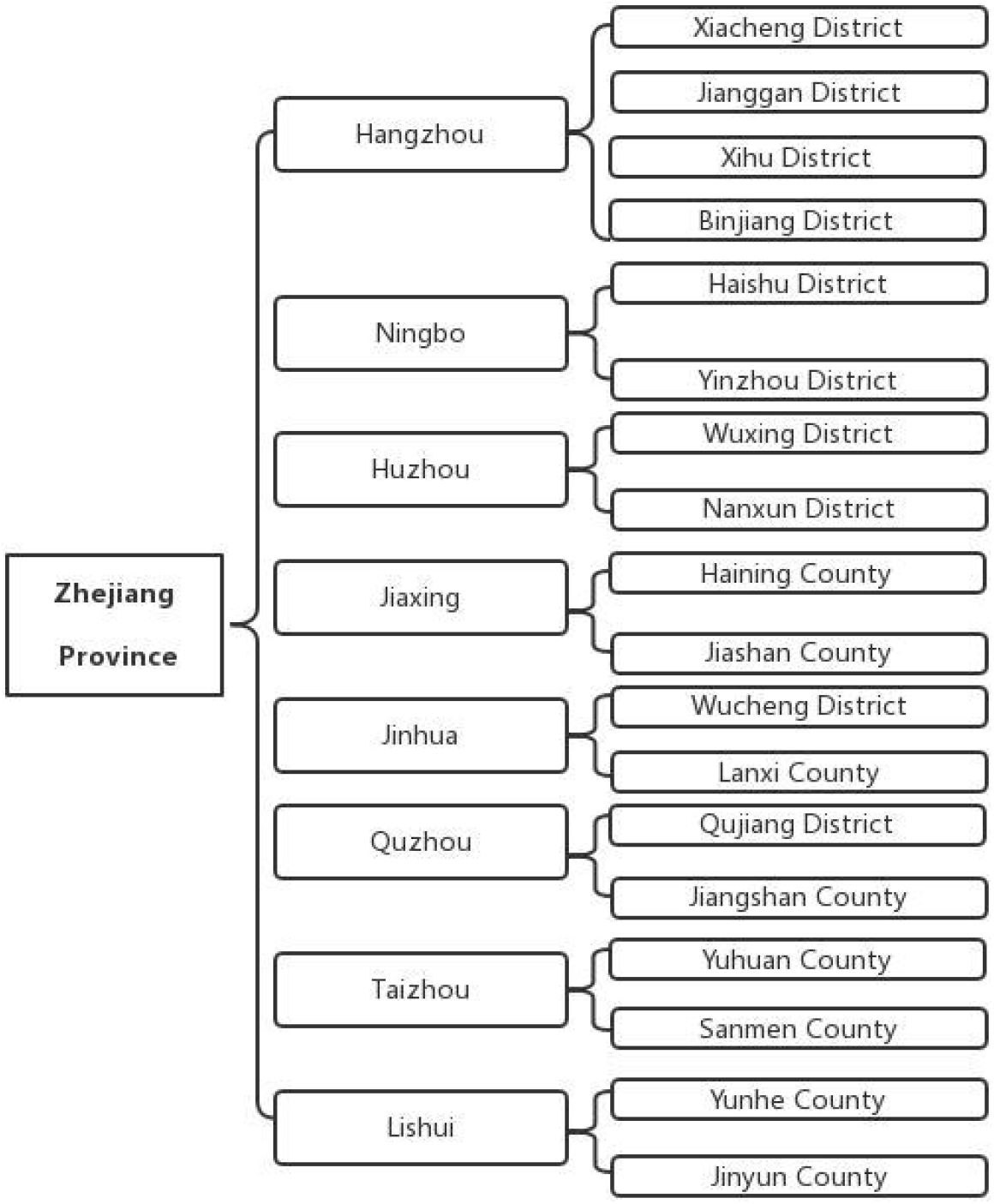
Scheme for sampling in this study. a. Eight selected cities in Zhengjiang province b. In each district/county, 1-2 street (township) community health service centers (CHSCs) were selected, from which about 100 guardians and 50 elderly were surveyed.

Guardians of 0–59-month-old children were randomly invited to participate in the study and questioned when they brought their children to the CHSC for health checkups or vaccinations. All questionnaires were completed by the guardians under the direction of trained interviewers. To ensure that the respondent was the decision-maker for the child’s vaccination, only the children’s parents were included.

The elderly aged ≥ 60 years who took free medical examinations in the CHSCs, which are required as part of the primary public health service in China, were randomly invited and included in the study. All questionnaires were completed by face-to-face interview by trained interviewers. The elders who failed to approach were excluded.

Based on the expected percentage of guardians having IVH of 50% and elders of IVH of 10%, with design effect of 3, the least sample size of guardians and elders was 1,160 and 420, respectively. Zhejiang Chinese Medical University Ethnics Committee reviewed and approved this protocol and informed consent was obtained from each subject.

## Data collection

The survey was conducted in the field, which comprised five sections: (i) demographic characteristics; (ii) IVH Scales for guardians (10 items) and elders (8 items): based on the ‘3C’ VH model proposed by SAGE^16^, and were modified after two rounds of expert Delphi consultation and preliminary survey; (iii) outcomes of future influenza vaccine intention to determine IVH, and influenza vaccination experience to determine IVD; (iv) 2 items ‘Have you heard of the influenza vaccine?’ and ‘Have you ever planned to (or bring your child to) get the influenza vaccination, but failed because of vaccine short supply?’ to ascertain the status of ‘not-knowing’ or ‘short supply;’ (v) items of related determinants for IVH, included from the SAGE VH determinants matrix^15^, including knowledge, experience, and influence from others and society for guardians, and an additional item of influenza vaccination policy for elders.

## Statistics analysis

Statistical analyses were performed in SPSS Version 25.0 (IBM Corporation, New York, NY, United States). To explore the structure of two scales, Exploratory Factor Analysis (EFA) was conducted on the samples using Principal Axis Factoring with orthogonal rotation (varimax). Cronbach’s α was calculated, and factors with Eigenvalues greater than one were extracted to determine internal consistency. The independent t-Sample test was used to evaluate the differences between two vaccine outcome questions to examine criterion validity of the scale. Binary Logistic regressions were used to explore related factors (demographic characteristics, knowledge, experience, influences from others and society, vaccine policy) of IVH and acceptance. According to the Hosmer–Lemeshow procedure, only covariates with p-value < 0.25 at the univariate analysis were entered into the multivariate analysis. The goodness of fit of the model was verified performing the Hosmer–Lemeshow test.

## Results

### Demographics

In total, 1,674 of the 1,800 guardians who were approached completed the interview, yielding a response rate of 93%. Of the 1,674 eligible participants, 71 (4%) were excluded from the analysis for failing to complete all items (44) or pass the quality control questions (27). For the elderly, 69% (553/800) were face-to-face interviewed and 31 (6%) were excluded from the analysis for inattentive or unmotivated responses.

The final sample in the analysis included 1,564 guardians and 522 elderly subjects. Of the guardians, 69.4% were mothers, 50% lived in an urban area, and 65.4% had a college education degree; with median age of 31 years. For the elderly, 58.4% were female, 56.7% lived in an urban area, and 6.7% had a college education degree; with the median age of 69 years (Table 1).

**Table 1.** Demographic characteristics of the guardians and the elderly included in the analysis

| Guardians (N=1564) |  | n (%) | The elderly (N=522) |  | n (%) |
| --- | --- | --- | --- | --- | --- |
| Gender |  |  |  |  |  |
|  | Male | 478 (30.6) | Male |  | 217(41.6) |
|  | Female | 1086 (69.4) | Female |  | 305(58.4) |
| Age |  |  |  |  |  |
| (year) | Range | 16-45 | Range |  | 60-92 |
|  | Median | 31 | Median |  | 69 |
| Ethnic |  |  |  |  |  |
|  | Han | 1514 (96.8) | Han |  | 518 (99.2) |
|  | Other | 50 (3.2) | Others |  | 4 (0.8) |
| Residen ce |  |  |  |  |  |
|  | Urban | 782 (50.0) | Urban |  | 296 (56.7) |
|  | Rural | 782 (50.0) | Rural |  | 226 (43.3) |
| Marital status |  |  |  |  |  |
|  | Married | 1553 (99.3) | Married |  | 414 (79.3) |
|  | Single/divorced | 11 (0.7) | Single/divorced /widowed |  | 108 (20.7) |
| Education |  |  |  |  |  |
|  | Middle school or lower | 250 (16.0) | Illiteracy |  | 97 (18.6) |
|  | High/secondary school | 292 (18.7) | Primary school |  | 181 (34.7) |
|  | Junior college | 310 (19.8) | Middle school |  | 149 (28.5) |
|  | Bachelor | 592 (37.9) | High/secondary school |  | 69 (13.2) |
|  | Master or higher | 120 (7.7) | College or higher |  | 26 (6.7) |
| Annual income (USD) |  |  |  |  |  |
|  | Decline to report | 89 (5.7) | Decline to report |  | 1 (0.2) |
|  | < 7016 | 169 (10.8) | < 2806 |  | 210 (40.2) |
|  | 7016-14031 | 321 (20.5) | 2806-7016 |  | 206 (39.5) |
|  | 14032-28063 | 434 (27.7) | 7017-14031 |  | 93 (17.8) |
|  | 28064-42093 | 261 (16.7) | ≥14032 |  | 12 (2.3) |
|  | 42094- | 290 (18.5) |  |  |  |
| Number of children |  |  | Live with their children |  |  |
|  | 1 | 863 (55.2) | Yes |  | 178 (34.1) |
|  | ≥2 | 701 (44.8) | No |  | 344 (65.9) |

| Age of child |  | Religion |  |
| --- | --- | --- | --- |
| < 6 months | 355 (22.7) | No | 345 (66.1) |
| 6-35 months | 877 (56.1) | Buddhism | 161 (30.8) |
| 36-59 months | 332 (21.2) | Others | 16 (3.1) |

### IVH and vaccine demand

In the 2018/2019 season, 51.9% (95% CI: 49.0–54.7%) of the guardians reported the influenza vaccination among children aged 6–59 months, and 9.6% (95% CI: 7.0–12.1%) of the elders also confirmed being vaccinated.

Fifty-one (4.2%) guardians and 262 (50.2%) elderly subjects had never heard of the influenza vaccine; only 4 (0.3%) guardians and 15 (2.9%) of elders completely rejected the vaccination; 522 (43.2%) guardians and 175 (33.5%) elders had IVH; 632 (52.3%) guardians and 70 (13.4%) elders completely accepted to be vaccinated, but only 379 (31.3%) and 29 (5.5%) of them had IVD (Figure 2).

**Figure 2.**
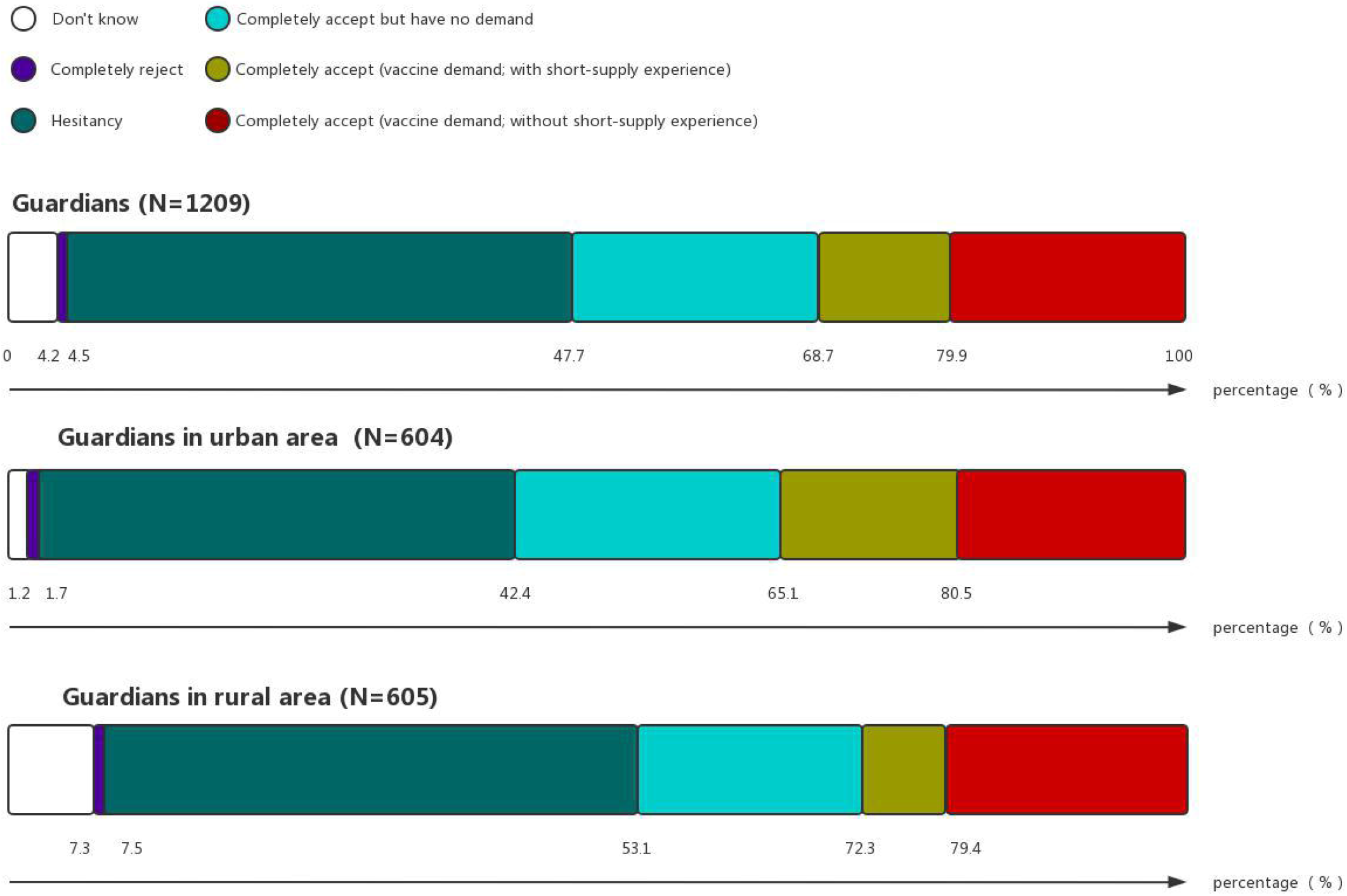
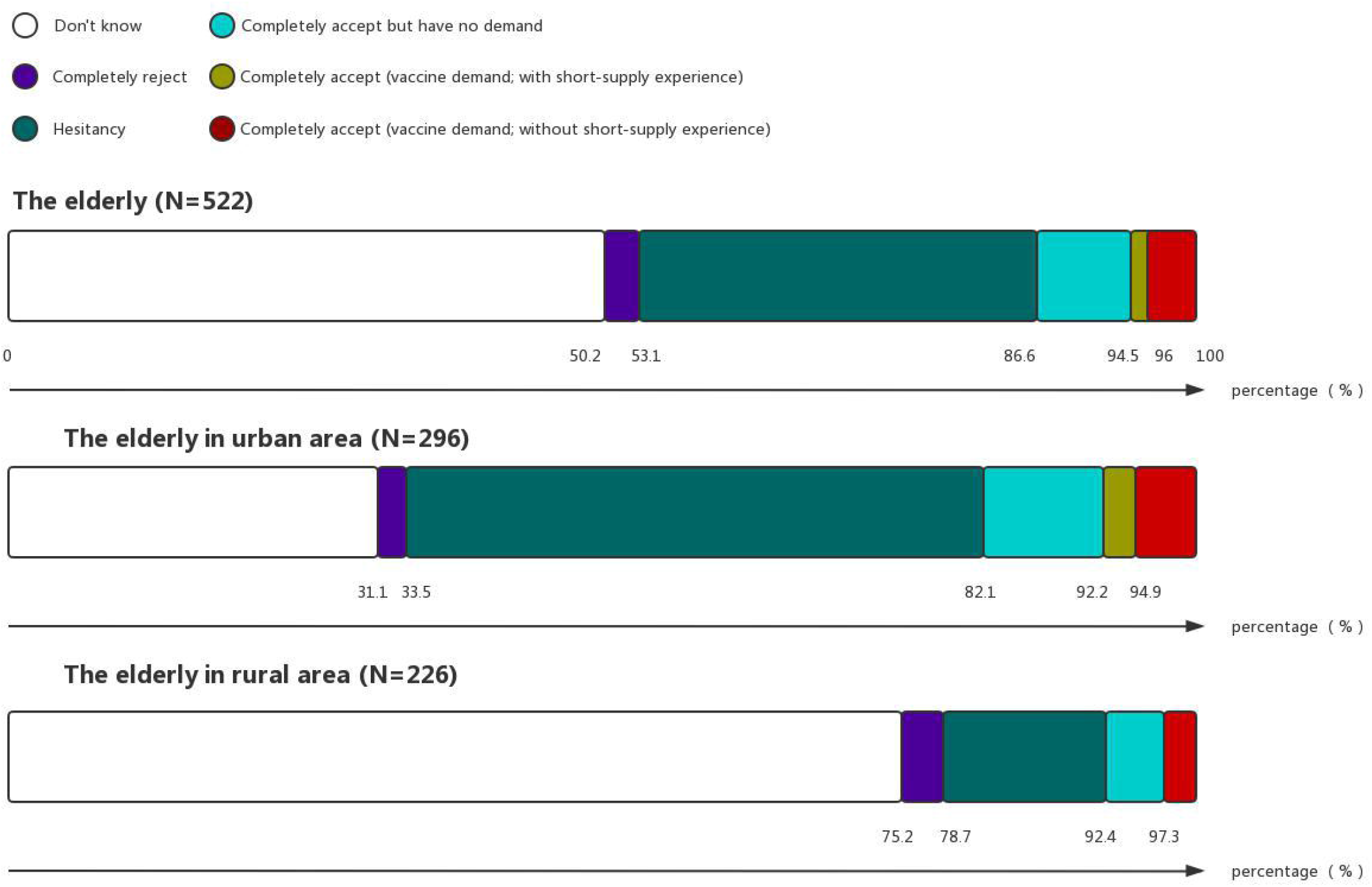
Distribution of influenza vaccine hesitancy and vaccine demand in guardians and the elderly. Note. Guardians whose infants were < 6 months old were excluded from the analysis, since they are not recommended for influenza vaccination.

### EFA of the IVH scales

For the guardians’ IVH scale, the KMO measure of sampling adequacy was 0.736, and Bartlett’s test of sphericity (p < 0.001) indicated that sufficient correlations among the variables existed, allowing to proceed. EFA identified four factors with Eigenvalues greater than one, explained 63.47% of the common variance of ten items. All the standardized loadings were > 0.6 and no cross loading was > 0.4, indicating that all items were significant. Finally, a reliability analysis revealed that the Cronbach’s α was 0.638.

For the elders’ IVH scale, it was shown that the KMO was 0.682with Bartlett’s test of sphericity, (p < 0.001). EFA identified three factors with Eigenvalues greater than one, explained 58.74% of the common variance of eight items. All the standardized loadings were greater than 0.6 and no cross loading was greater than 0.4. Reliability analysis revealed that the Cronbach’s α was 0.611.

Table 2 lists the items and results of EFA. Suppl Tab 1 shows linear correlations across dimensions.

**Table 2.**
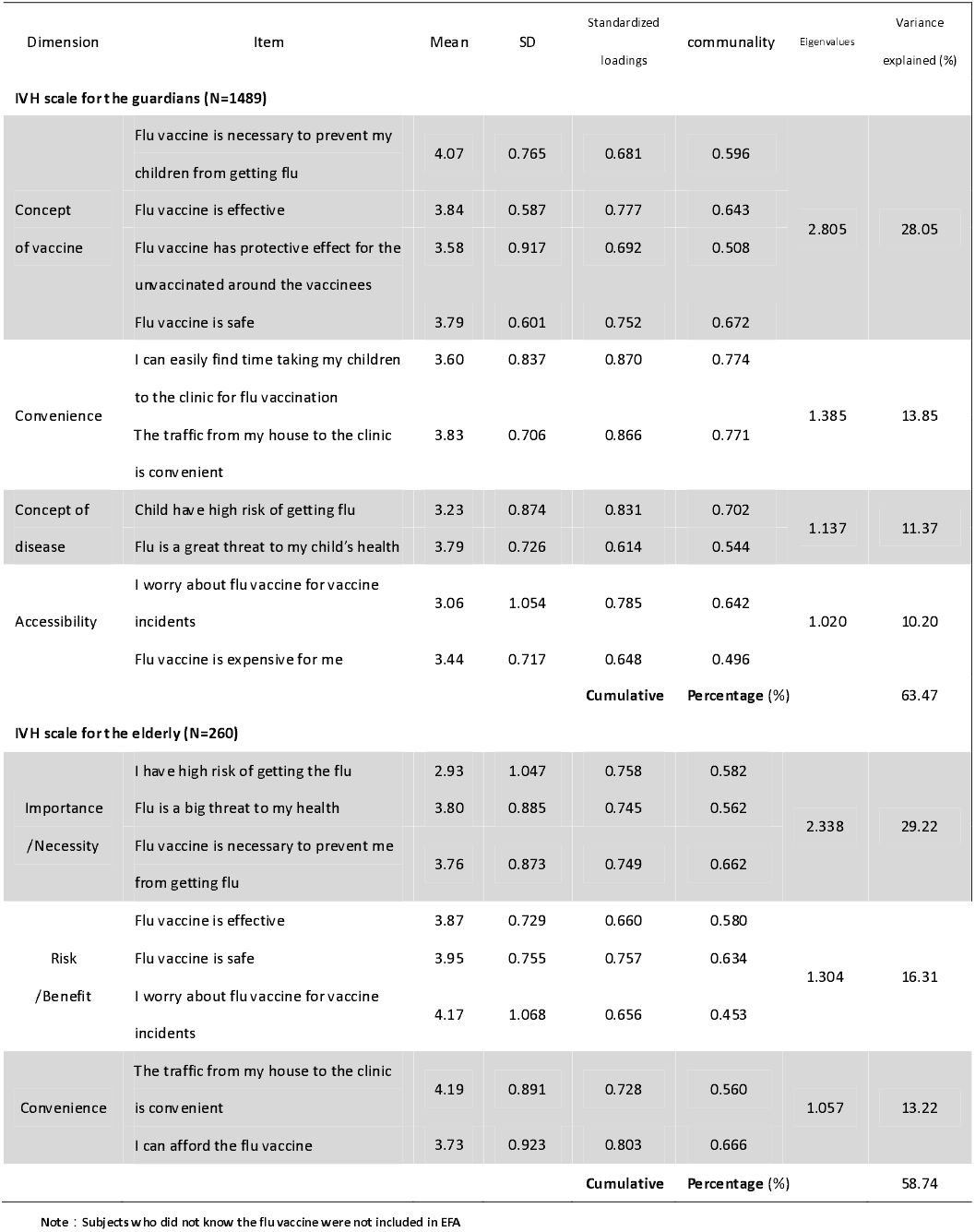
Exploratory factor analysis (EFA) for influenza vaccine hesitancy scales of the guardians and the elderly

### Criterion validity of the scales

The guardians who completely accepted vaccination had higher IVH score than those with hesitancy (Mean: 37.7 vs. 34.6, p < 0.001), and so did the elderly (Mean: 32.3 vs. 29.9, p < 0.001). Guardians who had received vaccination before reported higher IVH score than who had not the experience (Mean: 36.8 vs. 35.7, p < 0.001) and the same trend was noted in the elderly (Mean: 32.2 vs. 30.0, p < 0.001) (Table 3).

**Table 3.** Influenza vaccine hesitancy (IVH) scale scores between different groups of the subjects

|  | n | Mean | SD | t | p |
| --- | --- | --- | --- | --- | --- |
| **Guardians** |  |  |  |  |  |
| Future influenza vaccine intention (N=1484) |  |  |  |  |  |
| hesitant | 680 | 34.56 | 3.229 | 17.508 | <0.001 |
| Completely agree | 804 | 37.70 | 3.694 |  |  |
| Influenza vaccine experience (N=1158) |  |  |  |  |  |
| Never vaccinated | 531 | 35.69 | 3.740 | -4.786 | <0.001 |
| Vaccinated before | 627 | 36.77 | 3.918 |  |  |
| **The elderly** |  |  |  |  |  |
| Future influenza vaccination intention (N=245) |  |  |  |  |  |
| hesitant | 175 | 29.92 | 3.413 | 4.853 | <0.001 |
| Completely agree | 70 | 32.29 | 3.531 |  |  |
| Influenza vaccine experience (N=260) |  |  |  |  |  |
| Never vaccinated | 210 | 30.00 | 3.534 | -3.794 | <0.001 |
| Vaccinated before | 50 | 32.18 | 4.114 |  |  |

Note : (i) Subjects who had never heard of influenza vaccine was not included in the analysis.
(ii) The number of subjects who completely rejected influenza vaccine was small, so it was not included in the analysis.
(iii) The guardians of children aged < 6 months were not included in 'Past influenza vaccine experience for guardians.'

### Subgroups and characteristics of IVH

We used the median score as a cut-off value (guardians: 35, elders: 31) and classified the subjects with IVH into two groups: those with mild and severe IVH. Each item of scales was statistically different within the two groups (p < 0.001).

Of the ten items for guardians IVH, the ‘impact of vaccine incidents,’ ‘think children have a low probability of getting flu,’ and ‘the price of vaccine is too expensive’ were the most leading and important. For eight items in the elders questionnaire, the ‘think they have a low probability of getting flu,’ ‘think not necessary,’ and ‘the price of vaccine is too expensive’ were statistically different (Figure 3).

**Figure 3.**
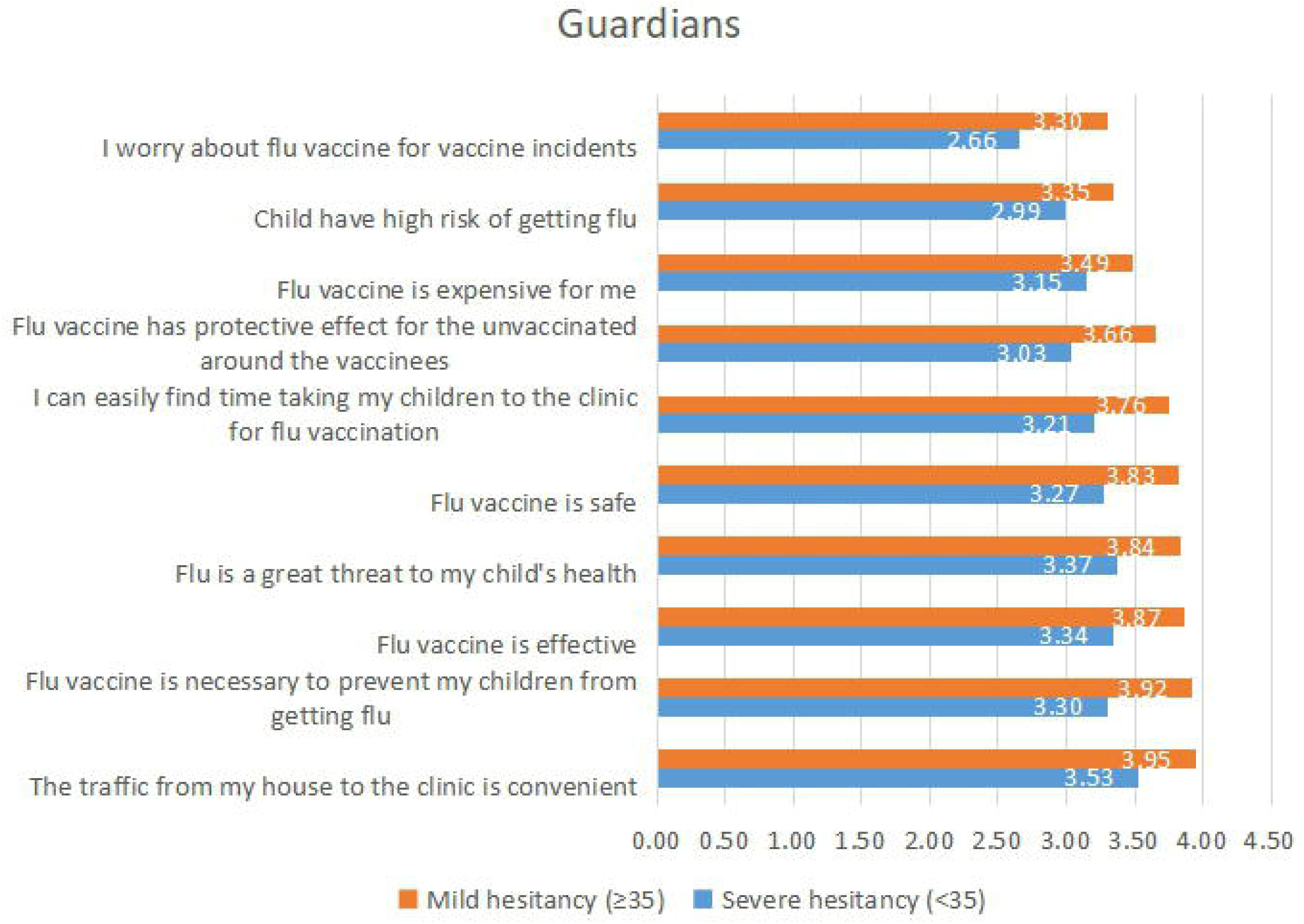
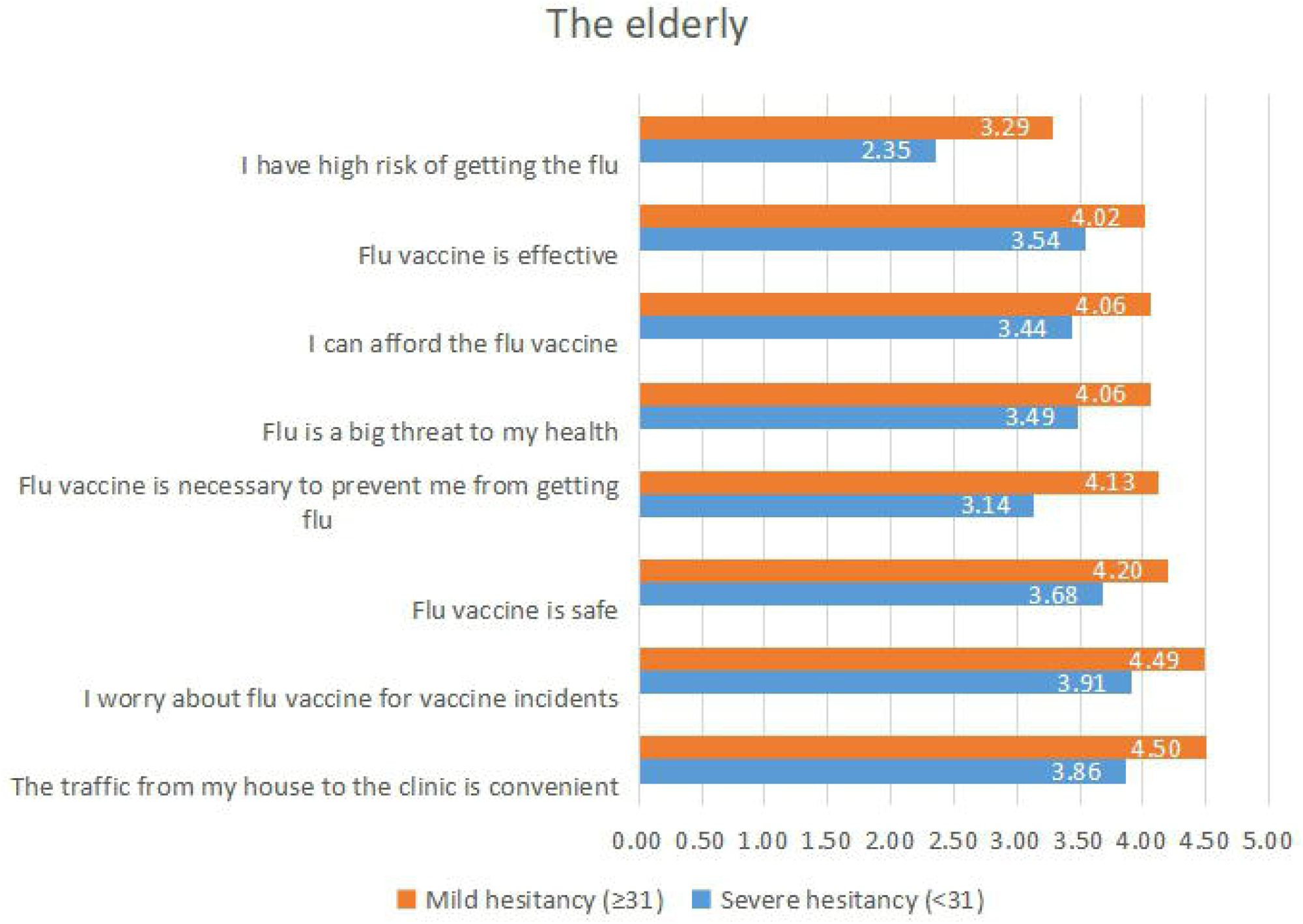
Subgroups and characteristics of influenza vaccine hesitancy (IVH) Note. Differences in each item between mild and severe hesitancy are significant (p <0.001) through independent sample t test.

### Determinants of VH

For guardians, variables of ‘from rural area’ (OR: 1.36, 95% CI: 1.03–1.79), ‘don’t know the government recommendation of flu vaccination’ (OR: 1.39, 95% CI: 1.06–1.83), and ‘don’t know flu vaccine is recommended to be vaccinated annually ‘(OR: 1.93, 95% CI: 1.47–2.53) were positively related with having IVH. The guardians whose family members (OR: 0.22, 95% CI: 0.16–0.29), or friends and neighbors (OR: 0.58, 95% CI: 0.44–0.76) had positive attitude toward vaccination were more likely to accept vaccination (Tab 4). Sensitivity analysis for guardians from urban or rural areas showed similar results (Suppl Tab 2).

The elderly, aged 70-79 years (OR: 0.46, 95% CI: 0.23–0.93), having influenza before (OR: 0.35, 95% CI: 0.16–0.77) or having vaccination experience (OR: 0.42, 95% CI: 0.18–0.98) were more likely to accept the influenza vaccination (Tab 5).

**Table 4.**
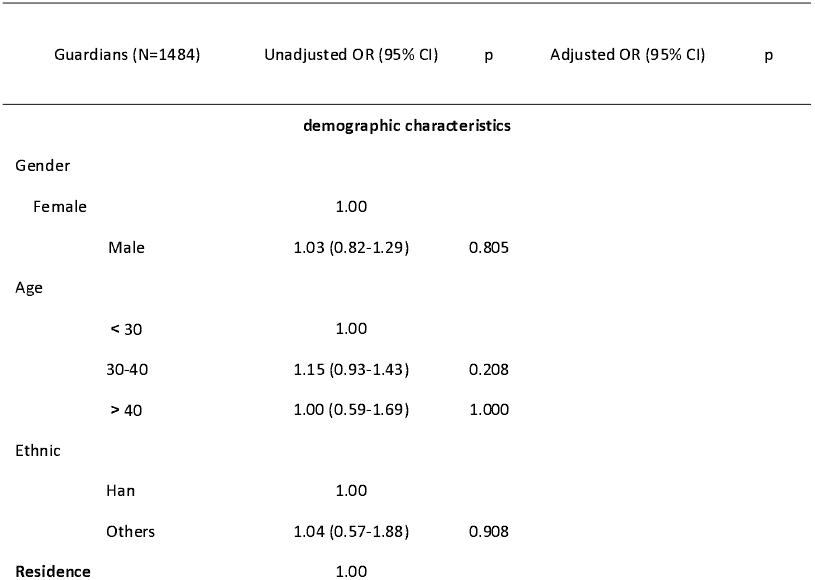
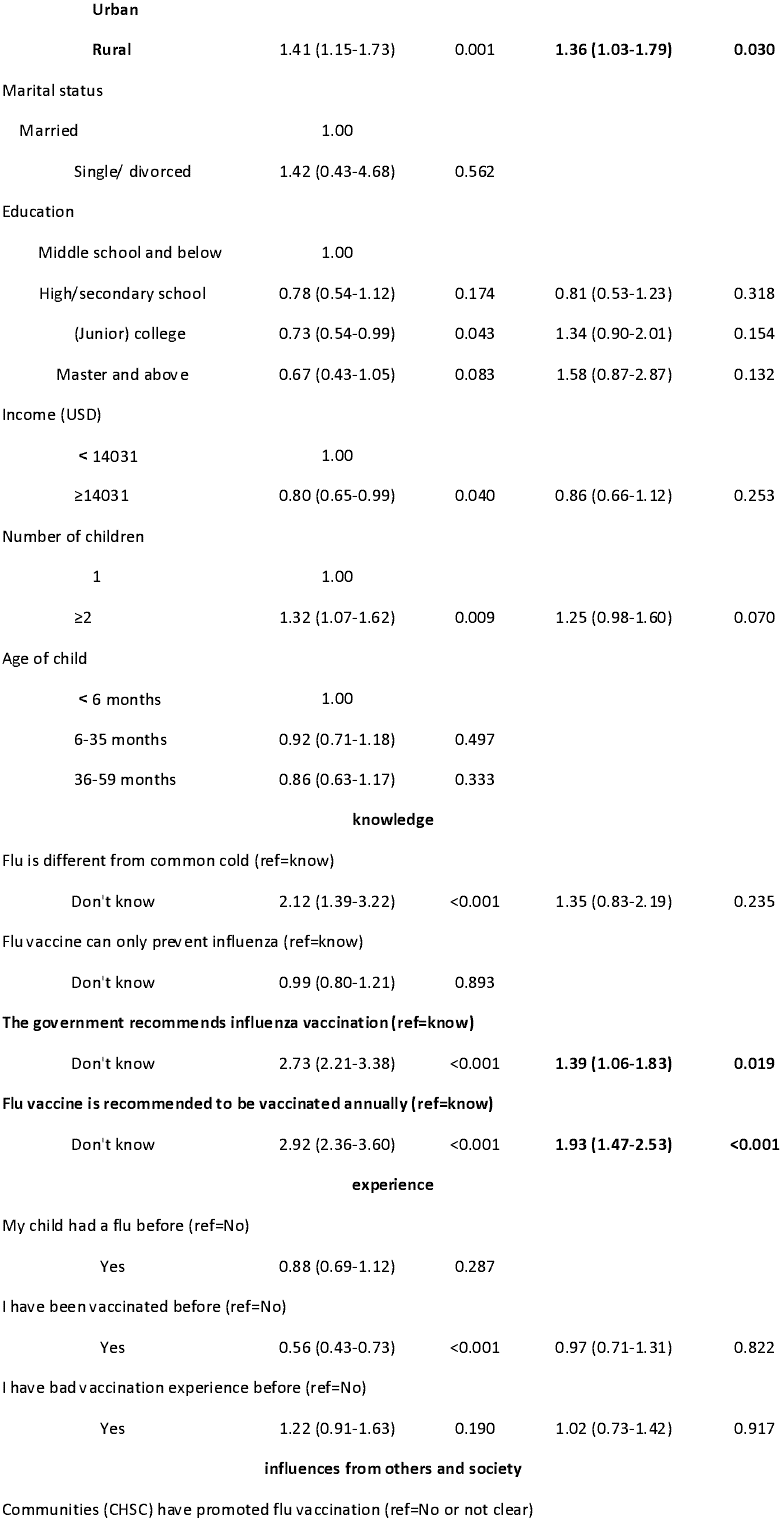
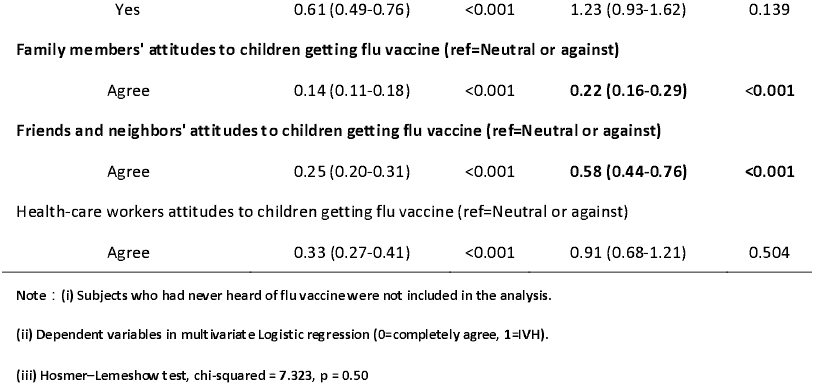
Determinants of influenza vaccine hesitancy in the guardians

| Guardians (N=1484) | Unadjusted OR (95% CI) | p | Adjusted OR (95% CI) | p |
| --- | --- | --- | --- | --- |
| **demographic characteristics** |  |  |  |  |
| **Gender** |  |  |  |  |
| Female | 1.00 |  |  |  |
| Male | 1.03 (0.82-1.29) | 0.805 |  |  |
| **Age** |  |  |  |  |
| < 30 | 1.00 |  |  |  |
| 30-40 | 1.15 (0.93-1.43) | 0.208 |  |  |
| > 40 | 1.00 (0.59-1.69) | 1.000 |  |  |
| **Ethnic** |  |  |  |  |
| Han | 1.00 |  |  |  |
| Others | 1.04 (0.57-1.88) | 0.908 |  |  |
| **Residence** |  |  |  |  |
|  | 1.00 |  |  |  |
| **Urban** |  |  |  |  |
| **Rural** | 1.41 (1.15-1.73) | 0.001 | **1.36 (1.03-1.79)** | **0.030** |
| **Marital status** |  |  |  |  |
| Married | 1.00 |  |  |  |
| Single/ divorced | 1.42 (0.43-4.68) | 0.562 |  |  |
| **Education** |  |  |  |  |
| Middle school and below | 1.00 |  |  |  |
| High/secondary school | 0.78 (0.54-1.12) | 0.174 | 0.81 (0.53-1.23) | 0.318 |
| (Junior) college | 0.73 (0.54-0.99) | 0.043 | 1.34 (0.90-2.01) | 0.154 |
| Master and above | 0.67 (0.43-1.05) | 0.083 | 1.58 (0.87-2.87) | 0.132 |
| **Income (USD)** |  |  |  |  |
| < 14031 | 1.00 |  |  |  |
| ≥14031 | 0.80 (0.65-0.99) | 0.040 | 0.86 (0.66-1.12) | 0.253 |
| **Number of children** |  |  |  |  |
| 1 | 1.00 |  |  |  |
| ≥2 | 1.32 (1.07-1.62) | 0.009 | 1.25 (0.98-1.60) | 0.070 |
| **Age of child** |  |  |  |  |
| < 6 months | 1.00 |  |  |  |
| 6-35 months | 0.92 (0.71-1.18) | 0.497 |  |  |
| 36-59 months | 0.86 (0.63-1.17) | 0.333 |  |  |
| **knowledge** |  |  |  |  |
| **Flu is different from common cold (ref=know)** |  |  |  |  |
| Don't know | 2.12 (1.39-3.22) | <0.001 | 1.35 (0.83-2.19) | 0.235 |
| **Flu vaccine can only prevent influenza (ref=know)** |  |  |  |  |
| Don't know | 0.99 (0.80-1.21) | 0.893 |  |  |
| **The government recommends influenza vaccination (ref=know)** |  |  |  |  |
| Don't know | 2.73 (2.21-3.38) | <0.001 | **1.39 (1.06-1.83)** | **0.019** |
| **Flu vaccine is recommended to be vaccinated annually (ref=know)** |  |  |  |  |
| Don't know | 2.92 (2.36-3.60) | <0.001 | **1.93 (1.47-2.53)** | **<0.001** |
| **experience** |  |  |  |  |
| **My child had a flu before (ref=No)** |  |  |  |  |
| Yes | 0.88 (0.69-1.12) | 0.287 |  |  |
| **I have been vaccinated before (ref=No)** |  |  |  |  |
| Yes | 0.56 (0.43-0.73) | <0.001 | 0.97 (0.71-1.31) | 0.822 |
| **I have bad vaccination experience before (ref=No)** |  |  |  |  |
| Yes | 1.22 (0.91-1.63) | 0.190 | 1.02 (0.73-1.42) | 0.917 |
| **influences from others and society** |  |  |  |  |
| **Communities (CHSC) have promoted flu vaccination (ref=No or not clear)** |  |  |  |  |
| Yes | 0.61 (0.49-0.76) | <0.001 | 1.23 (0.93-1.62) | 0.139 |
| **Family members' attitudes to children getting flu vaccine (ref=Neutral or against)** |  |  |  |  |
| Agree | 0.14 (0.11-0.18) | <0.001 | 0.22 (0.16-0.29) | <0.001 |
| **Friends and neighbors' attitudes to children getting flu vaccine (ref=Neutral or against)** |  |  |  |  |
| Agree | 0.25 (0.20-0.31) | <0.001 | 0.58 (0.44-0.76) | <0.001 |
| **Health-care workers attitudes to children getting flu vaccine (ref=Neutral or against)** |  |  |  |  |
| Agree | 0.33 (0.27-0.41) | <0.001 | 0.91 (0.68-1.21) | 0.504 |

Note : (i) Subjects who had never heard of flu vaccine were not included in the analysis.
(ii) Dependent variables in multivariate Logistic regression (0=completely agree, 1=IVH).
(iii) Hosmer–Lemeshow test, chi-squared = 7.323, p = 0.50

**Table 5.** Determinants of influenza vaccine hesitancy in the elderly

| The elderly ( N=245 ) | Unadjusted OR (95% CI) | p | Adjusted OR (95% CI) | p |
| --- | --- | --- | --- | --- |
| **Demographics** |  |  |  |  |
| **Gender** |  |  |  |  |
| Female | 1.00 |  |  |  |
| Male | 1.06 (0.60-1.87) | 0.836 |  |  |
| **Age** |  |  |  |  |
| 60-69 | 1.00 |  |  |  |
| 70-79 | 0.51 (0.28-0.91) | 0.024 | **0.46 (0.23-0.93)** | **0.030** |
| ≥80 | 0.61 (0.23-1.65) | 0.332 | 0.83 (0.23-3.06) | 0.784 |
| **Residence** |  |  |  |  |
| Urban | 1.00 |  |  |  |
| Rural | 0.67 (0.34-1.31) | 0.244 | 0.62 (0.24-1.55) | 0.303 |
| **Marital status** |  |  |  |  |
| Married | 1.00 |  |  |  |
| Single/Separated/Widowed | 0.63 (0.30-1.34) | 0.228 |  |  |
| **Live with children** |  |  |  |  |
| Yes | 1.00 |  |  |  |
| No | 1.16 (0.64-2.10) | 0.630 |  |  |
| **Education** |  |  |  |  |
| Primary school and below | 1.00 |  |  |  |
| Middle school | 1.52 (0.80-2.90) | 0.201 | 1.40 (0.63-3.09) | 0.411 |
| High/secondary school | 1.68 (0.76-3.70) | 0.196 | 1.25 (0.47-3.36) | 0.659 |
| (Junior) college and above | 2.18 (0.67-7.12) | 0.196 | 2.10 (0.45-9.89) | 0.349 |
| **Income (USD)** |  |  |  |  |
| < 7016 | 1.00 |  |  |  |
| ≥7016 | 0.70 (0.37-1.32) | 0.269 | 0.84 (0.37-1.92) | 0.684 |
| **Religion** |  |  |  |  |
| No religious affiliation | 1.00 |  |  |  |
| Buddhism | 0.77 (0.43-1.37) | 0.372 |  |  |
| **knowledge** |  |  |  |  |
| Flu is different from common cold (ref=know) |  |  |  |  |
| Don't know | 1.06 (0.50-2.27) | 0.870 |  |  |
| Flu vaccine can only prevent influenza (ref=know) |  |  |  |  |
| Don't know | 1.38 (0.79-2.41) | 0.259 |  |  |
| The government recommends influenza vaccination (ref=know) |  |  |  |  |
| Don't know | 1.12 (0.64-1.96) | 0.684 |  |  |
| Flu vaccine is suggested to be vaccinated every year (ref=know) |  |  |  |  |
| Don't know | 2.97 (1.67-5.29) | <0.001 | 1.65 (0.81-3.36) | 0.170 |
| **experience** |  |  |  |  |
| **Had a flu before (ref=No)** |  |  |  |  |
| Yes | 0.27 (0.14-0.53) | <0.001 | **0.35 (0.16-0.77)** | **0.009** |
| **Had been vaccinated before (ref=No)** |  |  |  |  |
| Yes | 0.18 (0.09-0.36) | <0.001 | **0.42 (0.18-0.98)** | **0.045** |
| I have bad vaccination experience before (ref=No) |  |  |  |  |
| Yes | 1.50 (0.41-5.54) | 0.545 |  |  |
| **influences from others and society** |  |  |  |  |
| Communities (CHSC) have promoted flu vaccination (ref=No or not clear) |  |  |  |  |
| Yes | 0.55 (0.31-0.98) | 0.043 | 0.71 (0.31-1.63) | 0.416 |
| Family members' attitudes to the elderly getting flu vaccine (ref=Neutral or against) |  |  |  |  |
| Agree | 0.33 (0.18-0.58) | <0.001 | 0.80 (0.37-1.72) | 0.562 |
| Friends and neighbors' attitudes to the elderly getting flu vaccine (ref=Neutral or against) |  |  |  |  |
| Agree | 0.26 (0.13-0.50) | <0.001 | 0.53 (0.22-1.24) | 0.140 |
| HCWs' attitudes to the elderly getting flu vaccine (ref=Neutral or against) |  |  |  |  |
| Agree | 0.21 (0.10-0.42) | <0.001 | 0.44 (0.17-1.11) | 0.083 |
| **vaccine policy** |  |  |  |  |
| Local free flu vaccine policy (ref=No) |  |  |  |  |
| Yes | 0.35 (0.18-0.67) | 0.002 | 0.64 (0.26-1.58) | 0.335 |

Note : (i) Subjects who had never heard of flu vaccine was not included in the analysis.
(ii) Dependent variables in multivariate logistic regression (0=completely agree, 1=I/H).
(iii) Hosmer–Lemeshow test, chi-squared = 2.483, p = 0.963

## Discussion

Using cross-sectional data from the field and based on designed IVH scales from an eastern province in China, we found that over two-fifths of the children’s guardians and one-third of the elderly were hesitant with influenza vaccination in 2019. Accordingly, 31.3% of the guardians and 5.5% of the elderly had IVD. Poor awareness of seasonal influenza or vaccination, relatives’ negative attitude, and lacking recommendation of the government are statistically related to being hesitant.

We described the hesitancy as an intention/attitude between complete rejection and acceptance, different from the one by SAGE emphasizing the behaviors between vaccine delay and refusal. There are controversies whether VH is defined as an attitude or behavior^20^. A systematic review reported that vaccination behavior was the main outcome in most of the studies (377/470), while the intention/attitude to vaccinate against influenza was assessed in 100 of the 470 studies^18^. It needs to be highlighted that equivocation on the decision on whether to accept vaccination is the core issue^15^. The definition of VH is sometimes used interchangeably with concepts such as vaccine demand^23^, so we defined IVD to distinguish them. IVD was described as ‘the behavior of seeking influenza vaccination service’ in the study, following SAGE’s opinion that vaccine demand and hesitancy are not completely congruent as individuals may fully accept vaccination without hesitancy but may not think they need vaccination or a specific vaccine^16^. The concept of vaccine demand can also account for those who have been vaccinated according to the recommended schedule yet still had concerns about their decisions to receive vaccination^23^. In our study, 52.3% of guardians and 13.4% of the elderly completely accepted that vaccination for influenza is important; however, nearly 40% and 60% of them, respectively, did not have IVD, which will be at risk of influenza virus infection, indicating that challenges arise among those who fail to seek the vaccination service despite the full acceptance in China. In our study, VH was assessed by one item based on definition, similarly to studies made in Romania^25^, Canada^26^ and France^27^. Average scores of VHS were also used to assess VH in the USA^28^, e.g., indicating that one quarter of American parents had IVH.

Hesitancy profile and determinants are country- and context-specific globally. A systematic review showed that most Chinese studies defined VH as a problem of vaccine safety and vaccine incident response. Moreover, these studies hold the government accountable because of problems related to regulation deficits and inappropriate crisis management^22^. Based on the ‘3 C’ model (complacency, confidence, and convenience) and previous studies^29,30,31,32^, we designed two scales to recognize the profiles of IVH among the guardians and the elderly in the country, and both passed the reliability and validity test. The three topmost serious issues (with the lowest item score) were, for the guardians, the impact of vaccine incidents, thinking that children have low probability of getting influenza, and the vaccine high cost. For the elderly, the challenges were thinking that they have a low probability of getting influenza, that vaccination is unnecessary, and the vaccine high cost. Hence, both guardians and elders thought the vaccine cost was too high for them and did not realize the necessity for influenza vaccination; same assumptions also appeared among health-care workers in China^33^. Globally, vaccine safety-related sentiment is particularly negative in the European region, while the Western Pacific region reported the highest level of religious incompatibility with vaccines^34^. A large-scale retrospective analysis also found that confidence in the importance of vaccines had the strongest association with vaccine uptake^35^. In our study, the self-reported influenza vaccine coverage in children and in elders was 52% and 9.6%, respectively. The former was higher than the national coverage (28.4%), while the latter was low and far away from the World Health Assembly target of 75%^36,37^. One half of the elders had never heard of the influenza vaccine; thus, the poor knowledge of the elderly is a huge challenge. Although nine tenths of the guardians had heard of influenza vaccine before, half of them did not know that the vaccine should be administered annually. Educational campaigns should be launched to improve guardians’ and elders’ awareness and knowledge of influenza vaccination^22^.

As vaccine safety has been one of the most important predictors of hesitancy worldwide, we, surprisingly, found that the guardians worried more about the unsafe (toxic/fake/expired) vaccines rather than adverse effects of the vaccine, which is the common worry in other countries^12^. The special contexts in China may lead to a scenario in which, i.e., vaccine incidents can affect vaccine confidence or coverage^15,38^ and we-media in internet may play a major role^19,39^. Like a double-edged sword, positive messages can help monitor VH^40^, increase knowledge, and boost vaccine coverage, while negative/false messages can affect vaccine confidence and lead to hesitancy, rejection, or an anti-vaccine sentiment^16,41^.

Short supply of influenza vaccine is a potential threat to the vaccine demand in China. Hesitancy may be present in situations where vaccine uptake is low because of lack of vaccine, stock-outs, or lack of vaccine offer. The SAGE Working Group agreed that these situations are not the principle driver of unvaccinated members of the population and fall outside the SAGE definition of VH^15^. The issue regarding short supply has become popular among those with IVD in China, i.e., 35.8% of the guardians and 27.3% of the elderly in our study reported their experience in short supply, which will increase the hesitancy and rejection of influenza vaccination as well as the increased infection risk. Stakeholders, including manufacturers and the government should evaluate the IVD in advance and provide the chance of getting vaccinated for those in the risk groups with the highest burden from the infection.

Determinants of IVH, including residence, knowledge of influenza and vaccine, recommendation from government, and the relatives’ attitude around (especially family members) can affect the guardian’s intention to vaccination. The elderly were more likely to be advised by the health-care workers in hospitals or communities, and those who had an influenza or vaccination experience were more willing to be vaccinated. Improving the knowledge of influenza and vaccines, as well as the support of relatives and the government are essential for deceasing IVH in Chinese guardians and elderly.

However, our study had several limitations. First, half of the elderly didn’t know about the influenza vaccine, as a result, the small sample size in the analysis may lead to the weak power in IVH and thus the generality may be hindered. Second, the samples were derived from one province in China, and thus the conclusions for IVH and IVD may not be generalized to other areas in the country. Third, the internal consistency of the two scales were up to standard but not fitted well enough, i.e., there were relatively less items in some dimensions and more items need to be explored and verified in future measurement.

## Conclusion

Over two-fifths of the guardians and one-third of the elderly were hesitant about influenza vaccination, while 31.3% of the guardians and 5.5% of the elderly had demand. Poor awareness of seasonal influenza and vaccination, relatives’ negative attitude, and the lack of government recommendations are statistically related to IVH. Worrying about poor quality of vaccines, and practical issues such as short supply of influenza vaccine may hinder the influenza vaccine confidence in China. More studies on hesitancy in populations with different cultural or historical backgrounds, and precision intervention to decrease the IVH in the guardian’s and elderly will be anticipated. Precision education aiming hesitancy in wider groups are anticipated to increase vaccine confidence and coverage in influenza-vulnerable groups.

## Data Availability

Data in this reasearch is all available

